# Social relationships and patient-reported outcomes in adolescent and young adult cancer survivors

**DOI:** 10.1101/2020.10.30.20223271

**Authors:** Pragya G. Poudel, Hailey E. Bauer, Zhaoming Wang, I-Chan Huang

## Abstract

**Importance:** Nearly 89,000 adolescents and young adults (AYAs) aged 15 to 39 years old are diagnosed with cancer in U.S. annually. Cancer diagnosis in AYAs often alters achievement of age-specific milestones, interferes with interpersonal relations, and disrupts social life. However, social relations in AYA survivors and associations with patient-reported outcomes (PROs) have been understudied.

**Objective:** To investigate the impact of cancer on PROs in AYA survivors and identify social integration mechanisms through which cancer experiences influence PROs.

**Design:** A cross-sectional study.

**Setting:** A national Internet survey panel maintained by Opinions 4 Good (Portsmouth, New Hampshire).

**Participants:** 102 AYA survivors and 102 age/sex/race-matched noncancer controls.

**Exposure:** Survivors were exposed to chemotherapy and/or radiotherapy during AYA.

**Main outcomes and measures:** Participants identified 25 closest friends/relatives they have contacted in past two years. Their interpersonal connections with each of 25 friends/relatives were used to create a social network index. The Duke-UNC Functional Social Support Questionnaire, UCLA Loneliness Scale, and PROMIS-29 Profile was used to measure social support, loneliness, and PROs (physical functioning, pain interference, fatigue, anxiety, and depression), respectively.

**Results:** AYA survivors of lymphoma, leukemia, and solid tumor had significantly better social networks than controls (all p-values <0.05). However, solid tumor and central nervous system malignancy survivors experienced higher loneliness than controls. Compared to controls, survivors had significantly poorer PROs in all domains. Cancer experience directly influenced all PRO domains (all p-values <0.05 except fatigue) and indirectly through social network-social support-loneliness pathways (all p-values <0.05). Survivors with high loneliness had lower physical functioning, higher pain interference, fatigue, anxiety, and depression compared with noncancer controls (all p-values <0.05).

**Conclusions and relevance:** AYA survivors were more socially connected, but experienced greater loneliness than controls. The perceived loneliness greatly influenced PROs. Future research should focus on the functional aspects of social relations rather than considering the structural aspects of social integration, which would provide an opportunity for appropriate interventions to improve health outcomes through social integration.

**KEY POINTS:** *Question:* How do social relationships associate with self-reported health outcomes between adolescent and young adult (AYA) cancer survivors and noncancer controls?

*Findings:* This cross-sectional study revealed that AYA survivors were more socially connected, but perceived greater loneliness compared to noncancer controls. AYA survivors with high loneliness had lower physical functioning, higher pain interference, fatigue, anxiety, and depression compared to noncancer controls.

*Meaning:* The findings of this study suggest that appropriate interventions, focused on improving functional social networks to further meet the needs of AYA cancer survivors, may function as a mean to prevent perceived loneliness and help achieve optimal health outcomes.

## INTRODUCTION

Nearly 89,000 adolescent and young adult (AYA) patients aged 15 to 39 years old are diagnosed with cancer in the United States annually.^1^ This is almost eight-times that of cancer diagnosed among ages 0 to 14 years, and five-times that of cancer diagnosed among age 40 and above.^1^ AYAs with cancer have significant decrements in physical and mental health status. In the AYA HOPE study, Smith et al. found poor self-reported physical and mental health in cancer survivors aged 25-34 and 35-44 years, respectively, compared with the general US population.^2^ Additionally, AYA cancer survivors are more likely to experience adverse clinical outcomes a few years after therapy completion, including obesity, cardiovascular disease, hypertension, and disability.^3, 4^ More importantly, AYAs are in a critical stage of developmental transition from childhood to adulthood.^5-7^ They face the challenges of completing education, pursuing employment, establishing economic independence from parents, finding a life partner, forming a family,^5, 6, 8, 9^ maintaining interpersonal relationships and managing social life.^10, 11^ However, cancer-related developmental and social outcomes in AYA survivors are understudied.^12, 13^

Prior studies have commonly reported survivors’ developmental challenges pertaining to academic achievement^14, 15^, employment opportunities^16, 17^, and finding a life partner^18^. A study based on the Childhood Cancer Survivor Study (CCSS) showed that of survivors diagnosed with cancer at an early stage of life, those treated for brain tumors, leukemia, and Hodgkin disease were more likely to receive special education compared with siblings.^19^ A review article identifies poor educational attainment, unemployment, and issues regarding interpersonal relationship as common issues in AYA cancer survivors. In particular, survivors with brain tumors were less likely to complete high school education as they are more likely to experience emotional health problems and physical disability than noncancer siblings.^20^ Along with poor educational attainment and unemployment, AYAs are less likely to be married and live independently compared to noncancer siblings. The risk for anxiety and depression remains high in cancer survivors.^21^

Social outcomes in cancer survivors have been widely identified through participants enrolled in the St. Jude Lifetime Cohort Study (SJLIFE).^22^ This study found hearing problems in survivors after cancer therapy which decreased social attainment. The study also showed that CNS tumor survivors in particular had poorer social attainment compared with non-CNS tumor survivors. Although these studies have explored social outcomes adhering to educational attainment, employment opportunities, marriage, and independent living, they did not evaluate social outcomes related to social integration and the subsequent impact on AYA cancer survivors. This study adopts the social integration model by Berkman et al. (Supplementary Table S1), including social network, social support and social isolation, to investigate the influence of these social integration variables on patient-reported outcomes (PROs) in AYA cancer survivors. ^23^

Structural ties or connections among people play a fundamental role to build social relations,^24^ which influence the health of an individual either directly and/or indirectly through social mechanisms including social support and perceived isolation. Factors such as sense of intimacy and connectedness provide stronger bonding which support for individual’s physical and mental well-being. In contrast, sense of social isolation, disconnectedness, and disintegration are linked to poor health outcomes.^23^ Evidence suggests that AYAs who

experience cancer at an early stage of life feel socially isolated^10, 25, 26^ as a result of treatment late effects that affect their interaction with and integration into society. Loneliness is a subjective perception of social isolation.^27, 28^ Lonelier individuals sense social threat and unsafe living in theirs environment. Feelings of loneliness accompanied by senses of anxiety, hostility, low self-confidence, and stress trigger behavioral and neurobiological mechanisms leading to adverse health consequences.^29, 30^ Such individuals may experience poorer physical functioning and greater depression, pain, and fatigue than socially connected individuals.^31^ Sense of social connectedness acts as an encouraging factor for an individual to adhere to positive health behaviors.^32, 33^

The objective of this study was to investigate social relationships and PROs between AYA cancer survivors and noncancer controls. We aimed to 1) compare three social relationship variables (social network, social support, and perceived loneliness) between survivors and controls, and 2) identify a specific mechanism among the aforementioned three social relationship variables through which cancer experience was associated with poor PROs.

## METHODS

### Study Design and Participants

This cross-sectional study included 102 AYA cancer survivors and 102 age/sex/race-matched noncancer controls. We recruited the participants from a national Internet survey panel maintained by Opinions 4 Good (Portsmouth, New Hampshire). The enrollment criteria for survivors were participants aged 18-30 years old at the time of the study, diagnosed with cancer at 15-30 years of age, and no cancer therapy in past three years. The enrollment criteria for controls were 18-30 years of age at the time of study and no history of cancer. This study was approved by the institutional review board of St. Jude Children’s Research Hospital in Memphis, Tennessee.

### Data collection

We performed data collection in spring of 2015. Participants who were eligible and agreed to participate in the study completed a self-administered survey via a secure website. We collected social network data using an egocentric approach. Participants were asked to identify 25 close friends and/or relatives with whom they frequently contacted in this past two years. They were then asked to indicate whether any of those friends and/or relatives knew each other, allowing us to create social tie data for up to 5,100 observations from 204 participants. Each participant’s identified friend and/or relative was asked about the type of relationship, type of communication used, and frequency of contact between themselves and the participant. The participants were also asked about the availability of resources for emotional support, tangible support, physical activity advice, and weight management advice from each of their friends/relatives. Using information regarding participant social ties, we created a functional social network index, with higher scores indicating better social network status.^34^

#### Measures

PROs were measured using the Patient-Reported Outcomes Measurement Information System - 29 Profile (PROMIS-29), a validated tool to measure generic PROs.^35^ We specifically focused on 5 PRO domains of interest: physical functioning, pain interference, fatigue, anxiety, and depression. Higher scores in the physical functioning domain indicated better PROs, while higher scores in other domains indicated worse PROs. Perceived social support was measured using the Duke-UNC Functional Social Support Questionnaire (8 items).^36^ High scores indicated better satisfaction with the quality of social integration with others. Loneliness was measured using the UCLA Loneliness Scale (20 items).^37^ High scores indicated more perceived disconnectedness.

Participants were asked about their sociodemographic characteristics including age, sex, race/ethnicity, educational attainment, annual household income (US dollar), and marital status. For the purpose of statistical analysis, we categorized sex as male and female; race/ethnicity as White, non-Hispanic, Black, non-Hispanic, Hispanic, and others; educational attainment as high school, college, and graduate levels; annual household income as <$40,000, $40,000 to $75,999, and ≥$80,000; and marital status as not married and married/living with partner as married.

### Statistical analysis

We performed Chi-square tests to test the difference in sociodemographic characteristics between survivors and controls. To compare social network, social support, and loneliness status between survivors and controls, we performed a linear regression analysis, adjusting for number of chronic health conditions. We used a path analytic model to quantify total, direct and indirect effects of cancer experience on each of the PRO domains through three social relationship variables (social network, social support and loneliness). Total effect was the sum of direct and indirect effects.

## RESULTS

Table 1 summarizes the characteristics of the study participants (N=204). In comparisons, all of the covariates were not statistically different between cancer survivors and noncancer controls (p-value >0.05), except marital status (p-value 0.046). Among survivors, 70% were diagnosed with cancer at age of 19-26 years and 30% at age of 15-18 years; 41.18% were treated for solid tumor, 26.47% for leukemia, 23.53% for Lymphoma, and 8.82% for central nervous system (CNS) malignancy. For cancer therapy, 61.76% and 29.41% of survivors were treated with chemotherapy and radiation therapy, respectively.

**Table 1:**
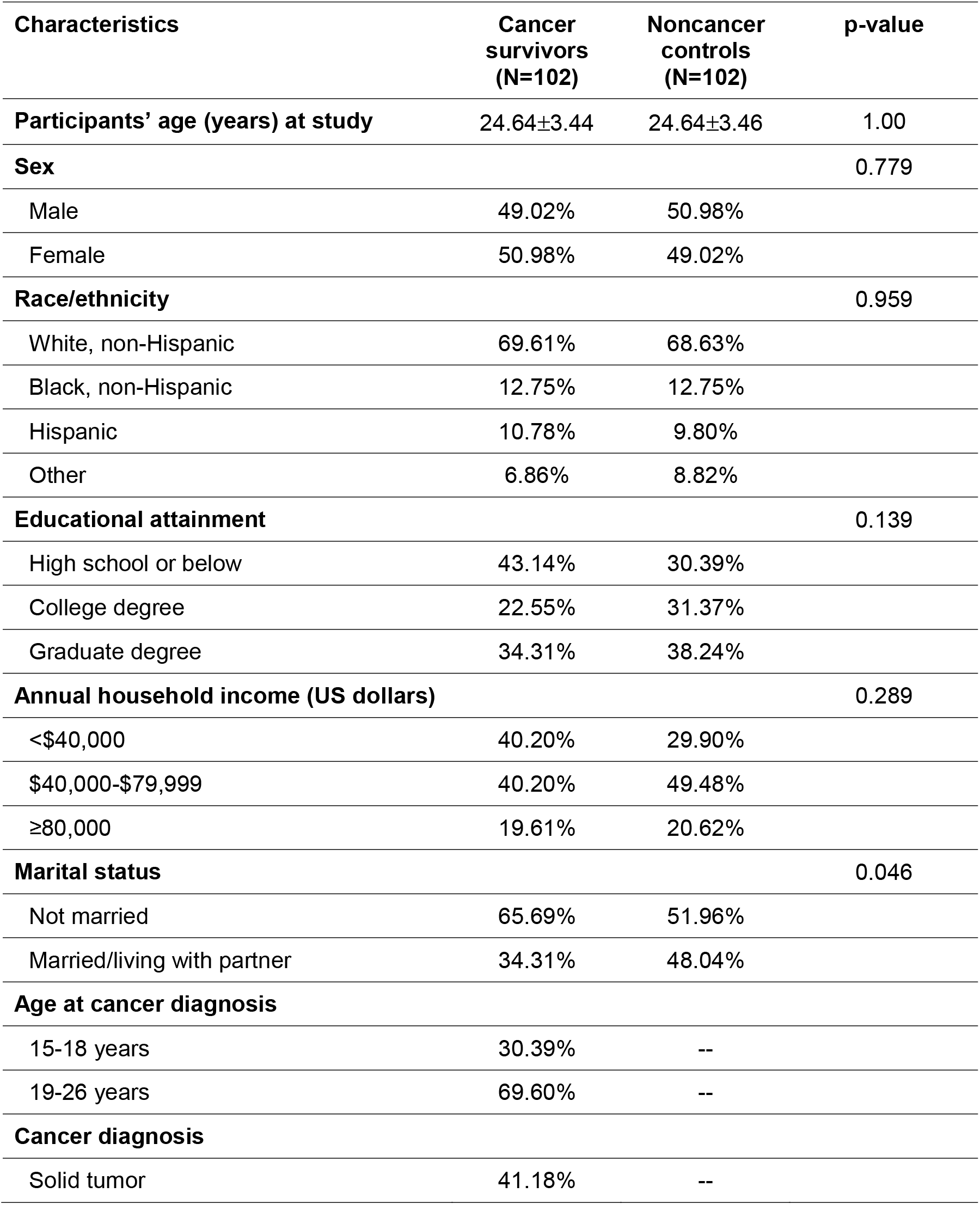
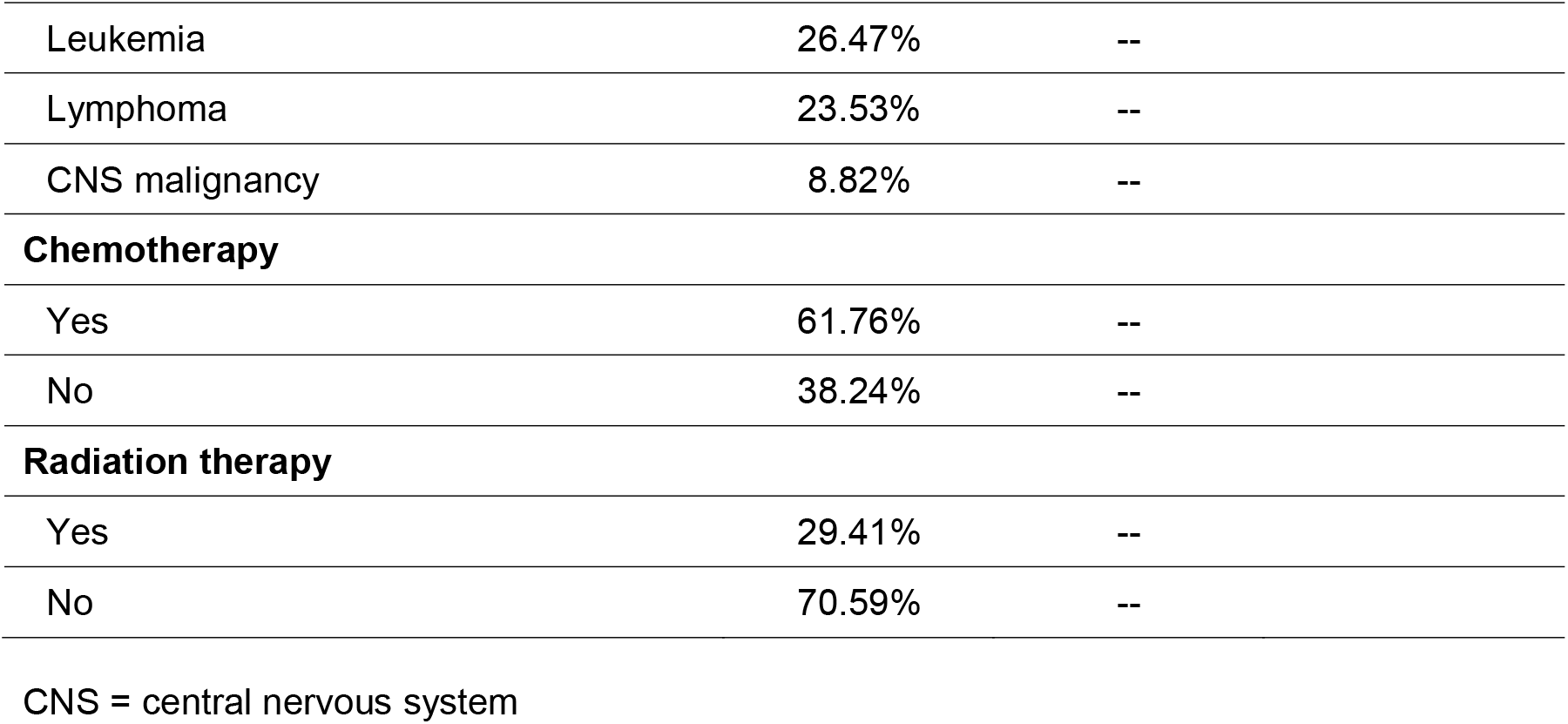
Characteristics of the study participants (N=204)

| Characteristics | Cancer survivors (N=102) | Noncancer controls (N=102) | p-value |
| --- | --- | --- | --- |
| **Participants' age (years) at study** | 24.64±3.44 | 24.64±3.46 | 1.00 |
| **Sex** |  |  | 0.779 |
| Male | 49.02% | 50.98% |  |
| Female | 50.98% | 49.02% |  |
| **Race/ethnicity** |  |  | 0.959 |
| White, non-Hispanic | 69.61% | 68.63% |  |
| Black, non-Hispanic | 12.75% | 12.75% |  |
| Hispanic | 10.78% | 9.80% |  |
| Other | 6.86% | 8.82% |  |
| **Educational attainment** |  |  | 0.139 |
| High school or below | 43.14% | 30.39% |  |
| College degree | 22.55% | 31.37% |  |
| Graduate degree | 34.31% | 38.24% |  |
| **Annual household income (US dollars)** |  |  | 0.289 |
| <\$40,000 | 40.20% | 29.90% | |
| \$40,000-\$79,999 | 40.20% | 49.48% | |
| ≥80,000 | 19.61% | 20.62% |  |
| **Marital status** |  |  | 0.046 |
| Not married | 65.69% | 51.96% |  |
| Married/living with partner | 34.31% | 48.04% |  |
| **Age at cancer diagnosis** |  |  |  |
| 15-18 years | 30.39% | -- |  |
| 19-26 years | 69.60% | -- |  |
| **Cancer diagnosis** |  |  |  |
| Solid tumor | 41.18% | -- |  |

|  |  |  |
| --- | --- | --- |
| Leukemia | 26.47% | -- |
| Lymphoma | 23.53% | -- |
| CNS malignancy | 8.82% | -- |
| **Chemotherapy** |  |  |
| Yes | 61.76% | -- |
| No | 38.24% | -- |
| **Radiation therapy** |  |  |
| Yes | 29.41% | -- |
| No | 70.59% | -- |

CNS = central nervous system

Table S1 presents social relationship variables between cancer survivors and noncancer controls with adjustments for the total number of chronic health conditions. Compared with controls, survivors of lymphoma, leukemia, and solid tumor had significantly higher functional social networks (B, 1.98, 95% CI, 0.60-3.36; B, 1.67; 95% CI, 0.38-2.96; B, 1.22; 95% CI, 0.12-2.33, respectively). However, solid tumor and CNS malignancy survivors experienced higher loneliness compared with controls (B, 10.83; 95% CI, 5.10, 16.57 and B, 15.65; 95% CI, 4.65-26.66). Experience of social support among lymphoma, leukemia, solid tumor and CNS malignancy survivors, and noncancer controls were comparable (p-value >0.05).

Table 2 and Figure 1 present the effects of cancer experience on individual PRO domains through the influence of three social relationship variables. For <u>physical functioning</u>, AYA cancer survivors reported significantly poorer physical functioning (B, -7.495; 95% CI, - 9.498 to -5.493) compared to controls. Approximately 84% of variance in poor physical functioning was directly influenced or explained by cancer experience (B, -6.333; 95% CI, - 8.201 to -4.464), whereas 16% of variance was explained through social relationship pathways. For <u>pain interference</u>, AYA cancer survivors had significant pain interference (B, 7.140; 95% CI, 3.665 to 8.472) compared to controls. However, 85% of variance in pain interference was directly influenced by cancer experience (B, 6.068; 95% CI, 3.665 to 8.472), whereas 15% of variance was explained through social relationship pathways. For <u>fatigue</u>, AYA cancer survivors reported significant fatigue (B, 4.542; 95% CI, 1.627 to 7.457) compared to controls. However, 40% of variance in fatigue was indirectly influenced through social relationship pathways. The significant pathway included poor social network to poor social support and to loneliness (B, 1.802; 95% CI, 0.216 to 3.338). For <u>anxiety</u>, AYA cancer survivors had significant anxiety (B, 6.015; 95% CI, 3.235 to 8.794) compared to controls. Approximately 70% of variance in anxiety was directly influenced by cancer experience (B, 6.015; 95% CI, 3.235 to 8.794) and 30% of variance was through social relationship pathways. The significant pathways included poor social network to poor social support and to loneliness (B, 1.905; 95% CI, 0.125 to 3.683). For <u>depression</u>, AYA cancer survivors had significant depression (B, 6.155; 95% CI, 3.542 to 8.768) compared to controls. Approximately 60% of variance in depression was directly influenced by cancer experience (B, 3.834; 95% CI, 1.745 to 5.923) and 40% of variance was through social relation pathways. The significant pathways included poor social network to poor social support and to loneliness (B, 2.321; 95% CI, 0.436 to 4.205).

**Table 2:**
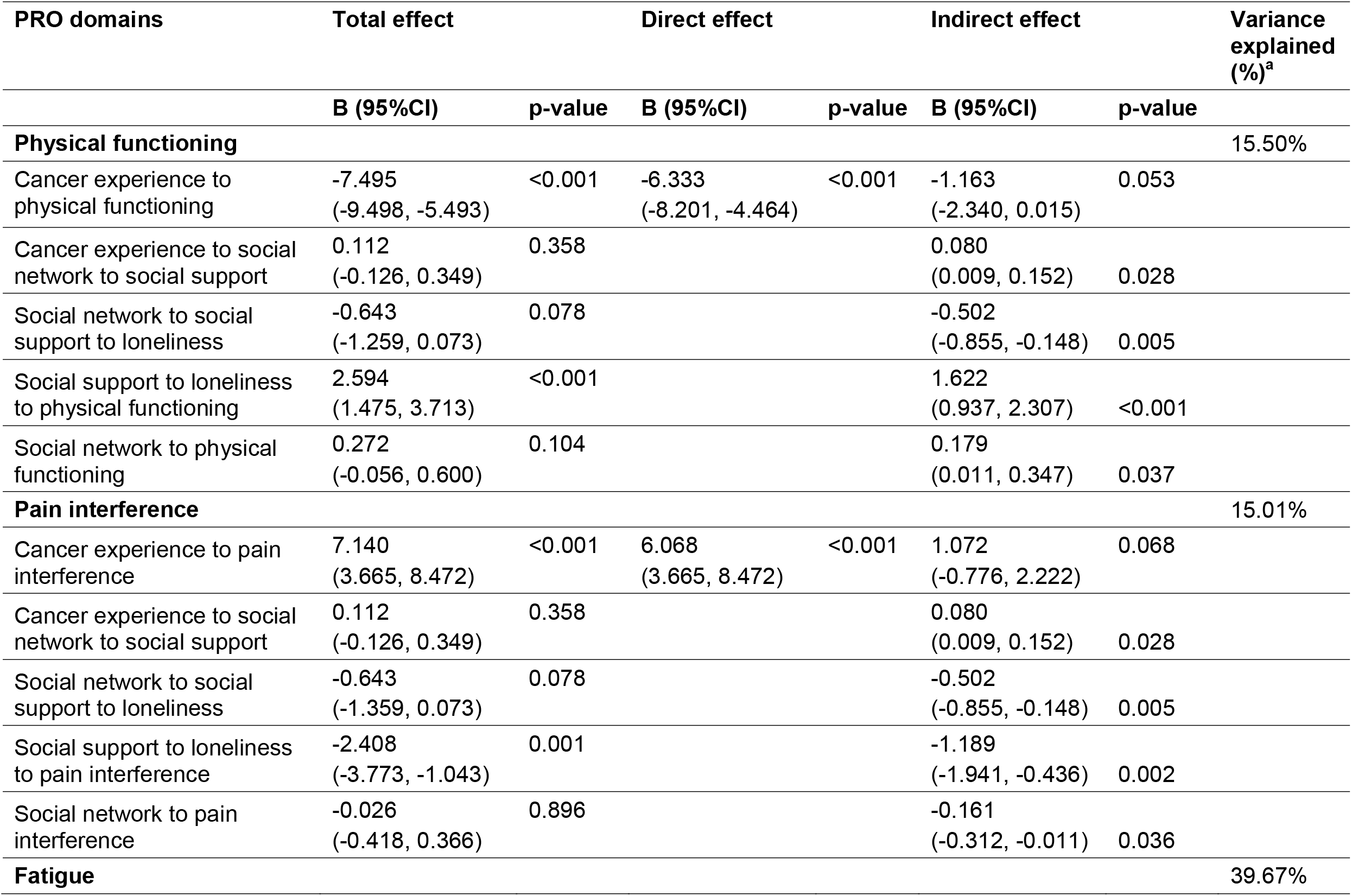
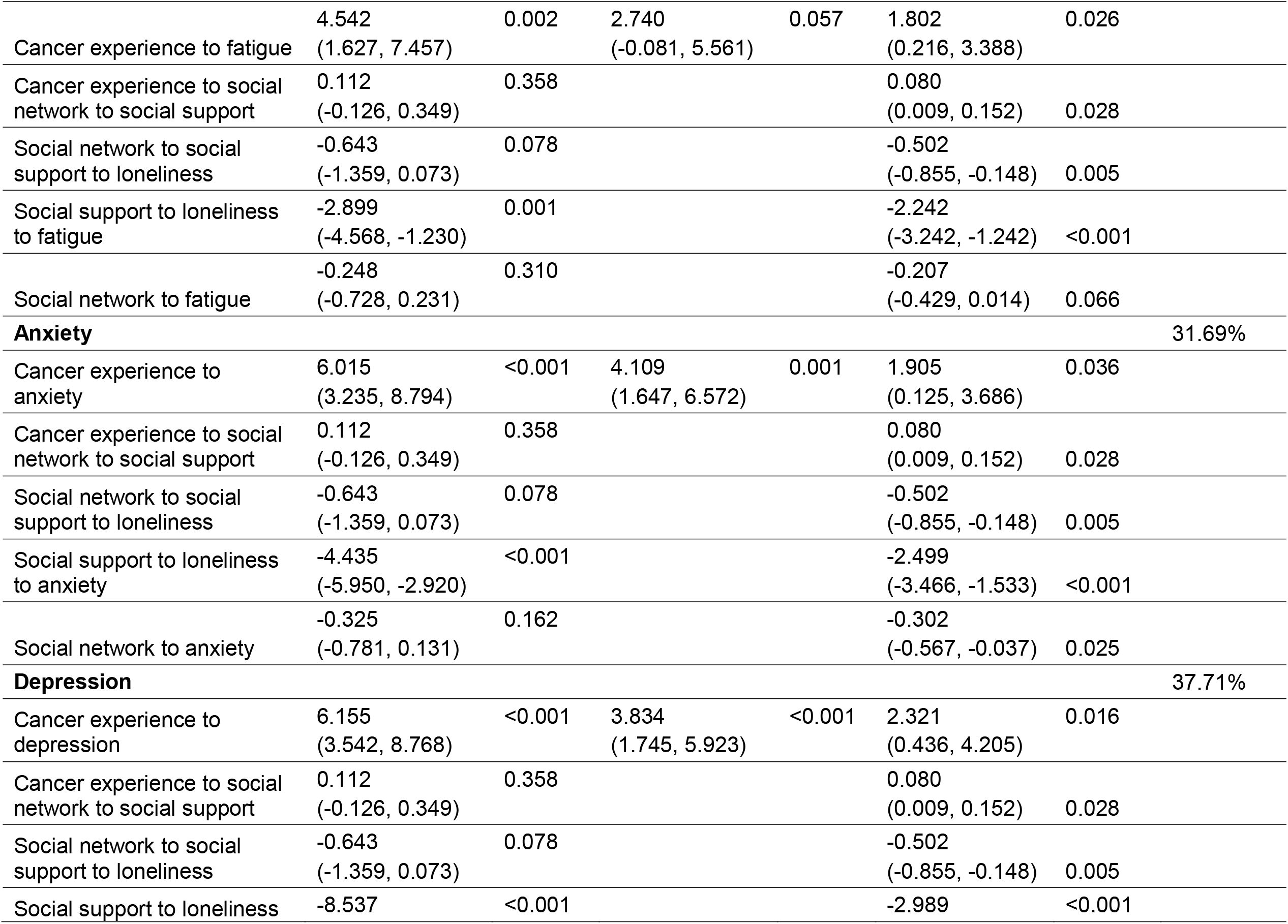
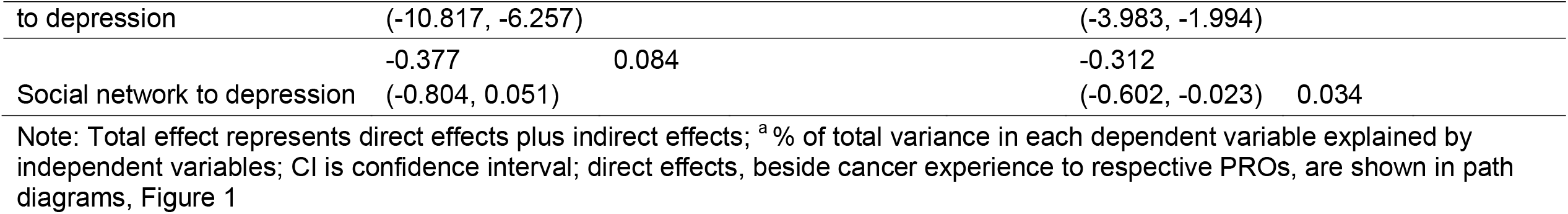
Effects of cancer experience on patient-reported outcomes (PROs) through social variables

**Figure 1:**
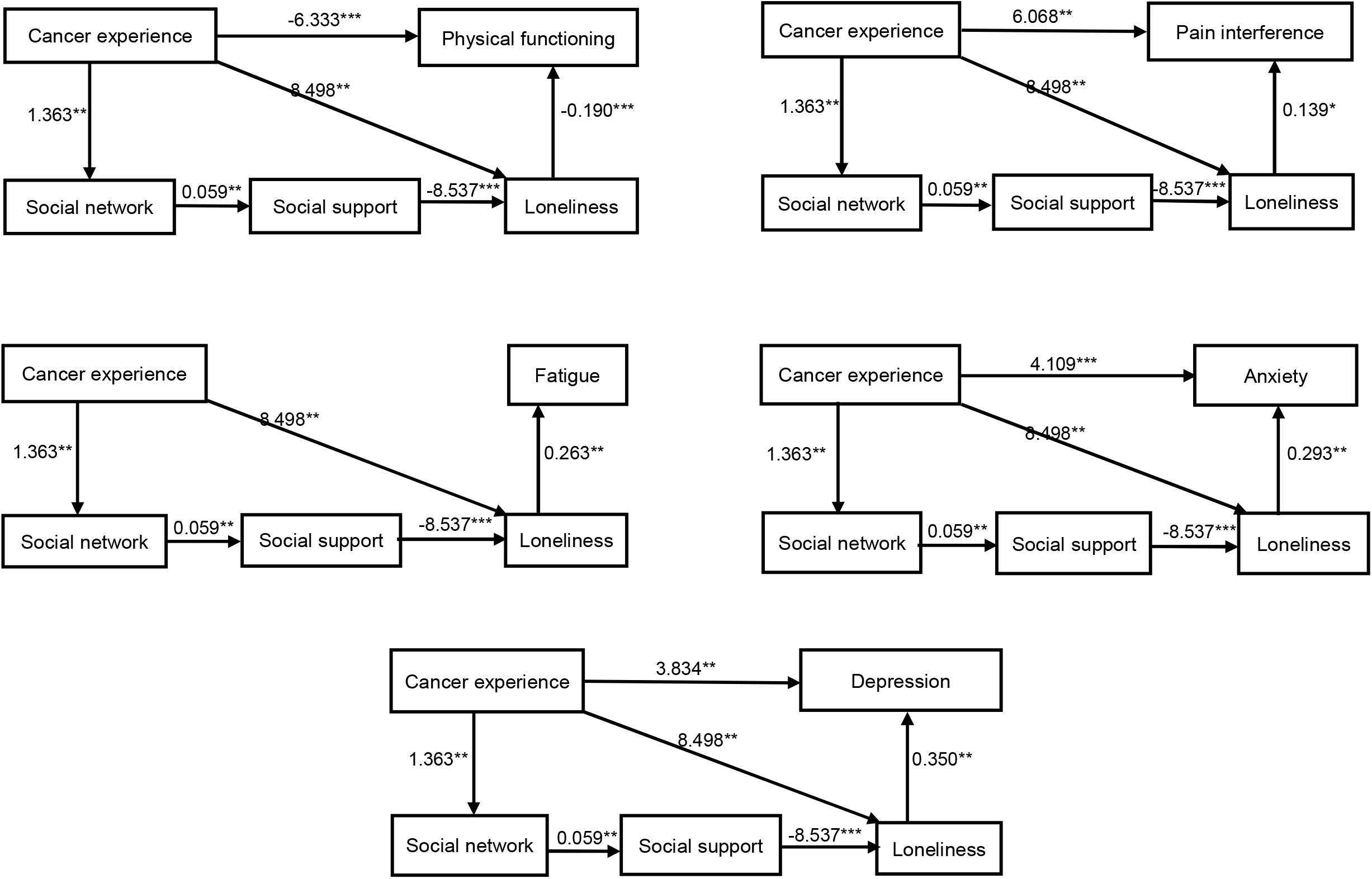
Effects of cancer experience on patient-reported outcomes through social variables. [Note: Values alongside the lines represent significant direct effects between variables; *p < 0.05; **p < 0.01; ***p < 0.001]

Table 3 presents associations of cancer experience and levels of loneliness with individual domains of PROs. Survivors having high loneliness had poorer PROs, typically in the physical, anxiety and depression domains, than controls having high loneliness, followed by survivors having low loneliness. Controls with low loneliness had the highest PROs. The magnitudes of decreased PROs in survivors with high loneliness in contrast to controls with low loneliness were approximately 2-fold the minimum important difference criterion, including depression (B, 12.20; 95% CI, 8.79 to 15.61), anxiety (B, 10.17; 95% CI, 6.29 to 14.04), poor physical functioning (B, -8.71; 95% CI, -11.42 to -6.00), pain interference (B, 7.38; 95% CI, 3.90 to 10.87), and fatigue (B, 6.82; 95% CI, 2.61 to 11.03).

**Table 3:** Cancer experience, loneliness, and patient reported outcomes.

| Cancer experience and loneliness | Patient-reported outcomes (PROs) |  |  |  |  |  |  |  |  |  |
| --- | --- | --- | --- | --- | --- | --- | --- | --- | --- | --- |
|  | Physical functioning |  | Pain interference |  | Fatigue |  | Anxiety |  | Depression |  |
|  | B (95%CI) | p-value | B (95%CI) | p-value | B (95%CI) | p-value | B (95%CI) | p-value | B (95%CI) | p-value |
| Controls with low loneliness | Ref |  | Ref |  | Ref |  | Ref |  | Ref |  |
| Survivors with low loneliness | -5.38<br>(-7.67, -3.09) | <0.001 | 4.94<br>(2.00, 7.88) | <0.01 | 0.50<br>(-3.05, 4.05) | 0.782 | 2.41<br>(-0.86, 5.68) | 0.147 | 2.32<br>(-0.55, 5.20) | 0.113 |
| Controls with high loneliness | -5.92<br>(-8.74, -3.11) | <0.001 | 3.49<br>(-0.13, 7.11) | 0.059 | 3.79<br>(-0.58, 8.15) | 0.089 | 7.40<br>(3.38, 11.42) | <0.001 | 9.06<br>(5.52, 12.60) | <0.001 |
| Survivors with high loneliness | -8.71<br>(-11.42, -6.00) | <0.001 | 7.38<br>(3.90, 10.87) | <0.001 | 6.82<br>(2.61, 11.03) | 0.002 | 10.17<br>(6.29, 14.04) | <0.001 | 12.20<br>(8.79, 15.61) | <0.001 |

Note: B-coefficient obtained after adjusting for age, sex, education, and total number of chronic health conditions; CI is confidence interval

## DISCUSSION

We found that AYA cancer survivors were more socially connected; however, they experienced more loneliness than noncancer controls. Specifically, survivors of solid tumor had more social connection, but solid tumor and CNS malignancy survivors experienced higher loneliness. Cancer experience had direct and indirect effects on PROs. Through the mechanism of three social relationship variables, survivors having better social connection were more socially supported and experienced lower loneliness, leading to better PROs of different domains.

Tremolada and colleagues explored health-related quality of life (HRQOL) and perceived social support in AYA pediatric cancer survivors. They found that survivors often lacked social support from family, friends, and significant others compared to controls, even if the survivors declared higher functioning on the social scale. The authors further explained that this could be because the survivors might have become more resilient to their situation over time. However, they did not explore survivors’ perception of social connectedness, which might have resulted in higher functioning on the social scale.^38^ Using an innovative design, our study compared social networks, social support, and loneliness simultaneously through a mechanistic aspect between AYA survivors and controls. Our study extended previous research that merely focused on the influence of personal social variables without considering social integration and interaction among individuals.

To identify meaningful psycho-social mechanisms that influence health outcomes in childhood cancer survivors, Huang et al. explored the mediational role of emotional distress on HRQOL in childhood cancer survivors using their siblings as the reference group. The types of emotional distress evaluated in that study included depression, anxiety, and somatization domains. The results revealed that survivors had poorer HRQOL than sibling controls. The path analysis further indicated the total effect of cancer experience on HRQOL was greatly explained by indirect effects through emotional distress. Additionally, cancer survivors experienced greater depression which was ultimately associated with greater role limitations due to physical health problems.^39^ This previous study investigated HRQOL through emotional distress while our current study explored PRO domains of physical functioning, pain interference, anxiety, fatigue, and depression as outcome variables through the influence of social integration in AYA survivors. Combined evidence from these two studies suggest that the influences of cancer experience on health status are complex, which can be through not only psychological but also social relationship routes.

A Previous stud identified loneliness as a significant risk factor for pain, depression, and fatigue among breast/colorectal cancer survivors.^40^ With the inclusion of noncancer controls, we found that AYA survivors with high loneliness were significantly associated with poor physical functioning, higher pain interference, fatigue, anxiety, and depression compared to controls with low loneliness. We further found that the controls with high loneliness experienced poor physical functioning, anxiety, and depression compared to controls with low loneliness. These findings suggest that experiencing loneliness, independent of cancer experience, might have a salient influence on poor PROs. Prevalence of perceived loneliness is increasing in our society across all age groups. We found that loneliness was significantly associated with poor health outcomes in AYA survivors, which was independent of influence from other social relationship variables. This suggests that individuals who perceive loneliness may not lack social relationships; instead, they perceive the relationships as poor quality. Loneliness is associated with poor physical and mental health consequences in adolescence and young adulthood.^41^ Interference of cancer or late medical effects in AYA regular activities may increase feeling of social isolation and concern for inability to achieve their life goals. Lower acceptance and support from peers are critical to social isolation.

The findings from this study provide useful implications for future clinical practice and research. It is important to assess related social integration factors during follow-up care as a part of risk assessment by primary care clinicians or survivorship clinicians who provide care to AYA survivors. As demonstrated in our study, AYA survivors experienced greater loneliness compared to controls, with better social networks associated with less loneliness. It is critical that the interventions address loneliness in order to achieve better health outcomes. Cheung and Zebrack found that AYA patients prefer resources that can mitigate the experience of isolation, build a sense of community or inclusive environment, and provide opportunities to connect with other AYA patients/survivors.^42^ In addition, AYA survivors are more inclined to connect with peer survivors as they share unique life challenges.^43^ Evidence suggests that interventions to share life experiences and build a sense of support system may help improve psychological well-being, self-efficacy and coping skills.^44^ Peer education has also helped in making healthy choices such as increase fruits and vegetables intake.^45^ A systematic review on health promotion and psychological interventions for AYA survivors revealed that peer-to-peer support was promising to achieve positive health outcomes.^46^ The interventions should take geographical, financial barriers, and cultural background of individuals into consideration.

In the digital age, AYAs are likely to benefit from the age-appropriate information provided through electronic applications.^6, 47, 48^ For example, a study that used Facebook-based intervention to increase physical activity among young adult survivors showed an increase in self-reported weekly minutes of moderate-to-vigorous physical activity.^49^ Use of health information available on the internet has increased confidence in patients byencouraging appropriate health-related decisions and asking questions to healthcare providers.^50^ However, use of technology-based interventions could be challenging when individuals lack technology skills or digital literacy, internet access, and sometimes even concern regarding the quality or accuracy of available medical related information.^51^ Thus, it would be appropriate to understand the population background and quality of available information before designing interventions.

### Limitations of the study

Despite of the innovative design to assess social integration and health outcomes in AYA cancer survivors, we acknowledge some limitations in our study. This is a cross-sectional study which precludes the provision of causal relationships; however, our social network measure was based on retrospective longitudinal data (i.e., experience in the past 2 years). Our study design relied on an egocentric approach. As a result, we were not able to investigate dynamic relationships between survivors and their friends/relatives. Future studies are warranted to focus on bidirectional interactions. Finally, we only evaluated PROs as proxies for objectively assessed late effects such as neurocognitive functioning and change of chronic health conditions. These late medical outcomes may be substantially impacted by social integration variables (e.g., social network and social isolation) and further investigation.

## CONCLUSIONS

We found that although AYA cancers have better social networks, they felt lonely, which in turn was associated with poor PROs. Thus, it is critical to use a social integration approach to address loneliness in order to achieve better health outcomes among AYA survivors. Future research should focus on functional aspects of social relations rather than considering structural dimensions or number of social connections and social ties. Screening social relationships in AYA cancer survivors and designing appropriate interventions to improve functional social integration should be an important element of follow-up care.

## Data Availability

We can provide data as per request.

