## Supplementary Tables and Figures for "Social relationships and patient-reported outcomes in adolescent and young adult cancer survivors"

**Supplementary materials**

Cancer experience

Social network

Social support

Loneliness

Patient reported outcomes

Figure S1: Conceptual framework showing the influence of social relationships on patient-reported outcomes

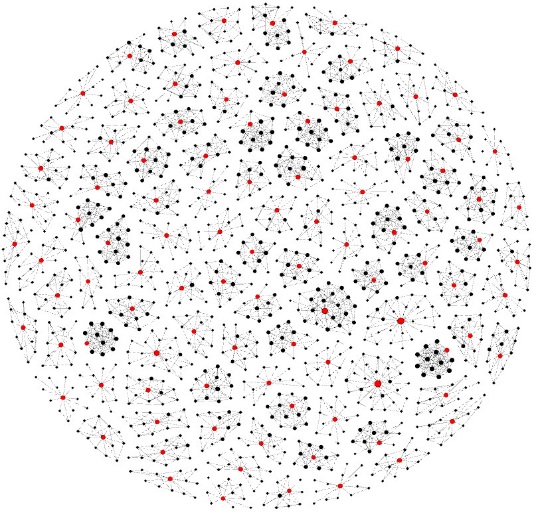

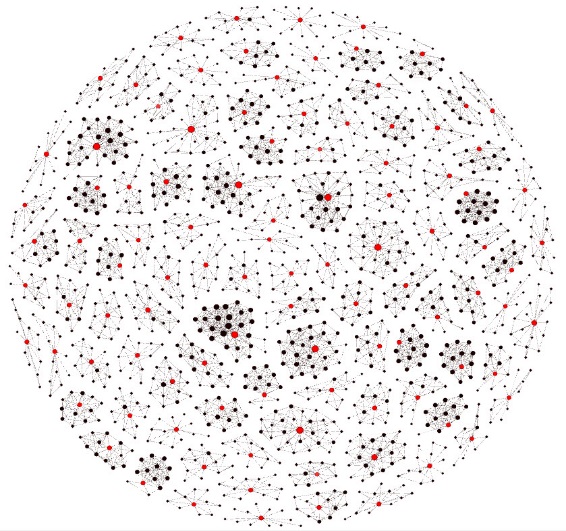

Survivors

Controls

Figure S2: Social connection map between survivors and controls

Table S1: Social network, social supports and loneliness by cancer survivors and noncancer controls

|  | **Social network** | | **Social support** | | **Loneliness** | |
| --- | --- | --- | --- | --- | --- | --- |
|  | **B**  **(95%CI)** | **p-value** | **B (95%CI)** | **p-value** | **B (95%CI)** | **p-value** |
| **Noncancer controls** | Ref | | Ref | | Ref | |
| **Cancer**  **survivors** | | | | | | |
| Lymphoma | 1.98  (0.60, 3.36) | 0.005 | 0.36  (-0.04, 0.75) | 0.078 | 0.05  (-7.10, 7.21) | 0.988 |
| Leukemia | 1.67  (0.38, 2.96) | 0.012 | 0.14  (-0.23, 0.51) | 0.453 | 5.85  (-0.86, 12.56) | 0.087 |
| Solid tumor | 1.22  0.12, 2.33) | 0.030 | 0.01  (-0.30, 0.33) | 0.931 | 10.83  (5.10, 16.57) | <0.01 |
| CNS malignancy | -1.09  (-3.21, 1.03) | 0.311 | -0.30  (-0.91, 0.31) | 0.333 | 15.65  (4.65, 26.66) | 0.006 |

Note: B-coefficient obtained after adjusting for total number of chronic health conditions; CI is confidence interval
